# Molecular Detection of Multi-drug resistant tuberculosis in clinical isolates from two urban centres in Malawi

**DOI:** 10.1101/2024.09.19.24313870

**Authors:** Tionge Sikwese, Takondwa Msosa, Hussein Twabi, Samuel Dzunda, David Chaima, Billy Banda, Yusuf Kanamazina, Mirriam Nyenje, Alinune Musopole, Marriott Nliwasa, Victor Ndhlovu

## Abstract

**Introduction:** Suboptimal chemotherapy allows *Mycobacterium tuberculosis* to develop drug resistance owing to development of resistant mutants in the mycobacterial population. Early diagnosis of TB and identification of drug-resistance is of particular importance in human immunodeficiency virus (HIV)-infected individuals, as delay of therapy and subsequent development of drug-resistant TB can be devastating due to compromised immune systems.

**Methodology:** We conducted a cross-sectional evaluation study using presumptive *M. tuberculosis* positive clinical isolates at two urban sites in Malawi (Blantyre and Lilongwe) to assess the presence of mutant genes on first and second line TB drugs using Line Probe Assay (LPA) and the gold standard drug susceptibility test (DST)

**Results:** For the Lilongwe site, the incidence of MDR-TB by Line Probe Assay (LPA) was found to be 14.06% (95% CI: 8%-20%) whereas that for Rif mono-resistance was 6.25% (95% CI: 2%-10%). Contrastingly, MDR-TB by DST was 23.44% (95 CI:16% - 21%) while mono-resistance was 6.25% (95% CI:2% -10). There was a substantial agreement on the detection of MDR-TB (kappa statistic was 0.75 with 95% CI of 0.62-0.88). Blantyre site, at 9.5% confidence interval, the point estimate for MDR-TB was 0% while for INH mono-resistance TB was 3.3%.

**Conclusions:** There is high incidence of MDR-TB among patients whose samples are sent to the Lilongwe site than previously thought. A short turnaround time to diagnosis, and the ability to simultaneously detect rifampicin and isoniazid resistance, makes LPA a reliable tool for the early detection of multidrug-resistant tuberculosis.

## Introduction

Tuberculosis (TB) remains a global public health emergency which until coronavirus (Covid19) was the leading cause of death from a single infectious agent *Mycobacterium tuberculosis* (*Mtb*) ranking above HIV/ AIDS[1]. Globally, there were 1.4 million TB deaths among HIV-negative people and an additional 187 000 among HIV-positive people in 2021 alone [1]. Suboptimal chemotherapy allows *M. tuberculosis* to develop drug resistance owing to development of resistant mutants which get introduced into the mycobacterial population [2,3]. Mono-resistant *M. tuberculosis* is when an isolate is resistant to only one drug, whereas multi-drug resistant (MDR) *M. tuberculosis* refers to the in-vitro resistance to at least rifampicin (RIF) and isoniazid (INH), the two main first line anti-TB drugs. A majority of RIF resistant *M. tuberculosis* isolates possess genetic mutations within the 81bp core region of the *rpoB* gene (Rifampicin resistance determining region RRDR) [4–6] whereas INH resistance is mediated via mutations within the *katG* gene or the promotor region of the *inhA* gene [7,8].

Multi-drug-resistant TB is an emerging issue in Malawi and results from the national TB prevalence survey completed in 2014 showed MDR-TB prevalence at 0.4% among new patients and 4.8 % in previously treated TB patient populations respectively [9].

Early diagnosis of TB and identification of drug-resistance is of particular significance in human immunodeficiency virus (HIV)-infected individuals, as delay of therapy and the resulting drug-resistant TB can be devastating especially for compromised immune systems.

Currently, phenotypic drug susceptibility testing (DST) remains the gold standard for the detection of resistance among TB patients. However, the method suffers from a long culture period ∼ 3-6 weeks to obtain results [10]. Additionally, critical drug concentrations for second-line drugs have not been completely established. The development of a new, rapid DST which is user friendly and inexpensive is thus an urgent priority for determining the susceptibility to second line drugs [11]. A rapid molecular test known as Geno Type MTBDR plus (Hain, Life Science) is a Line Probe Assay (LPA), which has been approved by WHO since 2008 for the diagnosis of MDR-TB [12], uses Polymerase Chain Reaction (PCR) and reverse hybridization methods for the rapid detection of mutations associated with drug resistance [13]. They are designed to identify Mycobacterium Tuberculosis Complex (MTBC) and simultaneously detect mutations in the *rpoB*, *katG* and *inhA* genes associated with drug resistance [14,15].

Patients infected with MDR-TB strains have particularly poor treatment outcomes and are more likely to remain sources of infection for longer periods of time [8,16,17]. This study was conducted to investigate the emerging cases of MDR-TB from two urban centres in Malawi (Blantyre and Lilongwe) and evaluate the performance of LPA against the standard DST. Monitoring circulating MDR-TB strains remains a critical part of any TB control program and a key factor to reduce and contain the spread of these resistant strains. Understanding the mechanisms of action of anti-TB drugs and the development of drug resistance will allow identifying new drug targets and better ways to detect drug resistance [5]. To date and to the best of our knowledge, few studies in Malawi have focused on the burden of MDR-TB using molecular based techniques [18,19]. Additionally, such studies are limited as they have focused only on particular geographical regions of the country and may not be representative of national data.

## Methodology

### Ethical consideration

Ethical clearance was obtained from the College of Medicine Research Ethics Committee (Certificate number COMREC: P.09/21/3418)

### Study setting and population

This was a cross-sectional evaluation study using pre-existing *M. tuberculosis* positive clinical isolates from Kamuzu University of Health Sciences (KUHeS)/Malawi Liverpool Welcome Trust (MLW) TB laboratory and the TB national Reference Laboratory complete with patient data. The study was approved by the College of Medicine Research Ethics Committee (Comrec). All the isolates originating from KUHeS/MLW TB laboratory belonged to a previous study (Exact TB study), with inclusion criteria: adults aged ≥ 18 years, having TB symptoms (cough of any duration, fever, night sweats or weight loss), able to submit sputum, willing and able to provide informed consent. Exclusion criteria: Presence of clinical danger signs (unable to walk unaided, confused or agitated, breathless when speaking or at rest), already taking TB treatment, or taken TB treatment within the last 60 days, unable to submit sputum and refuses offer of sputum induction, previously been enrolled in the trial), which was conducted at ART clinics in Bangwe Health Centre and Queen Elizabeth Central Hospital in Blantyre, Malawi. These health centres represent high density suburbs of Blantyre with estimated TB prevalence of 1,014 per 100,000. The KUHeS/MLW TB laboratory receives samples within Blantyre and surrounding areas, while the National TB reference laboratory in Lilongwe, receives samples from all the regions of Malawi which are suspected to be MDR-TB (These are patients who are already on TB treatment).

### Study procedures

The isolates were retrieved from frozen state using the unique patient identification number (PID) and generated laboratory number, allowed to thaw at 37°C before being inoculated into tubes of Middlebrook 7H9 broth base supplemented with oleic acid, albumin, dextrose and catalase (OADC) and an antibiotic mixture of polymyxinB, amphotericinB, nalidixicacid, trimethoprimandazlocillin (PANTA).The tubes were loaded in a BACTEC MGIT 96o machine 37 ◦C monitored weekly for growth of the culture.

Genotype MTBDRplus VER 2.0-line probe assay and Phenotypic DST Laboratory methods were used to detect MDR-TB in clinical *M. tuberculosis* positive clinical Isolates. Figure 1 below shows the simplified procedural flow diagram

**Figure 1.**
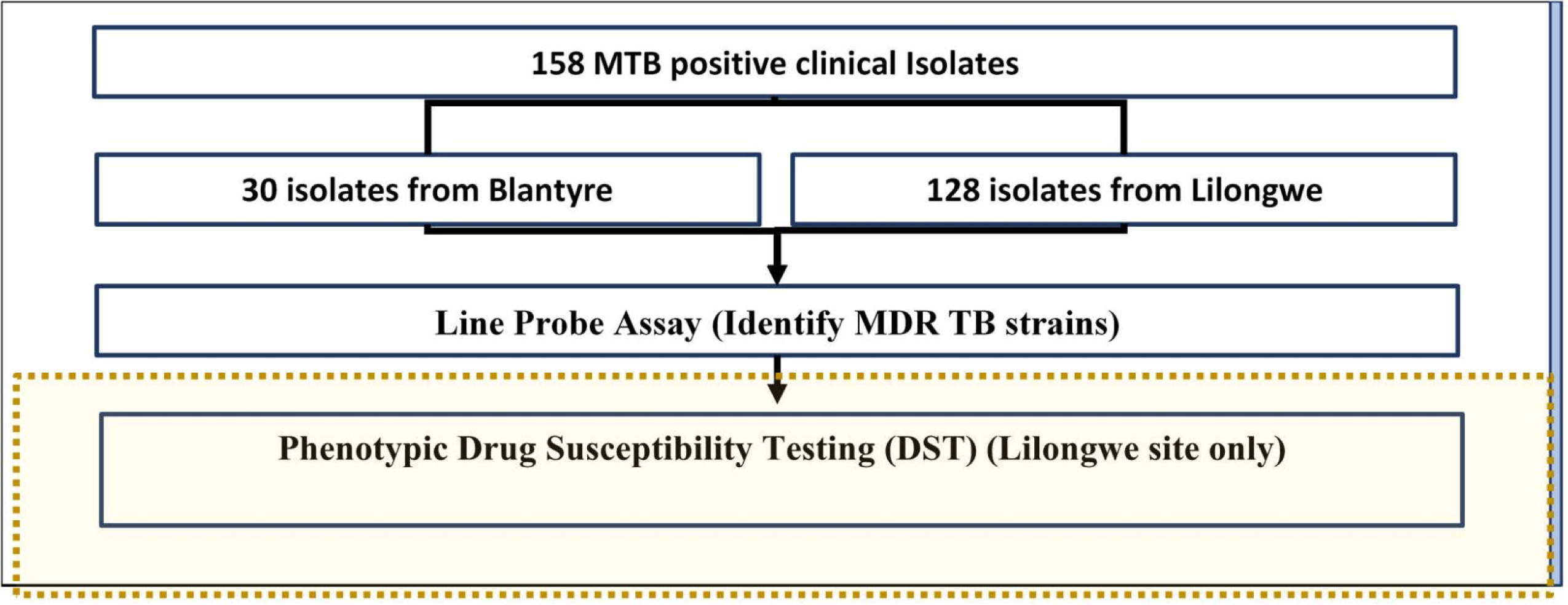
A flow chart of the laboratory methods; A total of 158 M. tuberculosis positive samples were used, 128 from Lilongwe and 30 from Blantyre. Samples from Lilongwe were subjected to both LPA and DST while those from Blantyre were only tested using LPA

### Line probe assay

The GenoType MTBDRplus VER 2.0 a DNA-strip-based in-vitro assay was performed according to the manufacturer’s (Hain Lifescience, Nehren, Germany) instructions. The steps for LPA assay included, DNA isolation, multiplex polymerase chain reaction (PCR) amplification and reverse hybridization. Mycobacterial DNA was extracted in BSL-3 laboratory following manufacturer’s instructions. In brief, a 500 ml aliquot of MGIT sample was centrifuged at 10,000Xg for 15 min, the supernatant was discarded and the pellet re-suspended in 100 ml sterile distilled water. The suspension was then heat killed at 85°C for 40 minutes on a heat block. The sample was sonicated for 15 min followed by centrifugation at 13,000 Xg for 10 minutes. A 5 ml aliquot of the DNA sample was used for PCR with the master mixture for amplification composing of 35 ml primer nucleotide mixture (provided with kit), 5 ml of 10XPCR buffer with 15 mM MgCl2, 2 ml of 25 mM MgCl_2_, 0.2 ml of HotStarTaq DNA polymerase (Hain Lifescience, Nehren, Germany), 3 ml nuclease free molecular grade water and 5 ml of DNA sample in a final volume of 50 ml. The amplification protocol consisted of 15 minutes of denaturation at 95°C, followed by 10 cycles of denaturation at 95°C for 30 seconds and 58°C for 2 min. This was followed by 20 cycles at 95°C for 25 seconds, 53°C for 40 seconds and 70°C for 40 seconds and a final extension at 70°C for 8 minutes. Hybridization was performed with the automatic machine (twincubator). After hybridization and washing, strips were removed, fixed on paper and the results were interpreted. The MTBDRplus assay strip contains 27 reaction zones; 21 of them are probes for mutations and six are control probes for verification of the test procedures. The six control probes included a conjugate control and amplification control (AC), an *M. tuberculosis* complex-specific control (TUB), an *rpoB* locus AC, a *katG* locus AC, and an *inhA* locus AC. For the detection of RIF resistance, the probes cover the *rpoB* gene, while for the INH resistance, specific probes cover positions in *katG* and *inhA*. The absence of at least one of the wild-type bands or the presence of bands indicating a mutation in each drug resistance-related gene implies that the sample tested is resistant to the respective antibiotic. When all the wild-type probes of a gene stain positive and there is no detectable mutation within the region examined, the sample tested is susceptible to the respective antibiotic (Figure 2).

**Figure 2.**
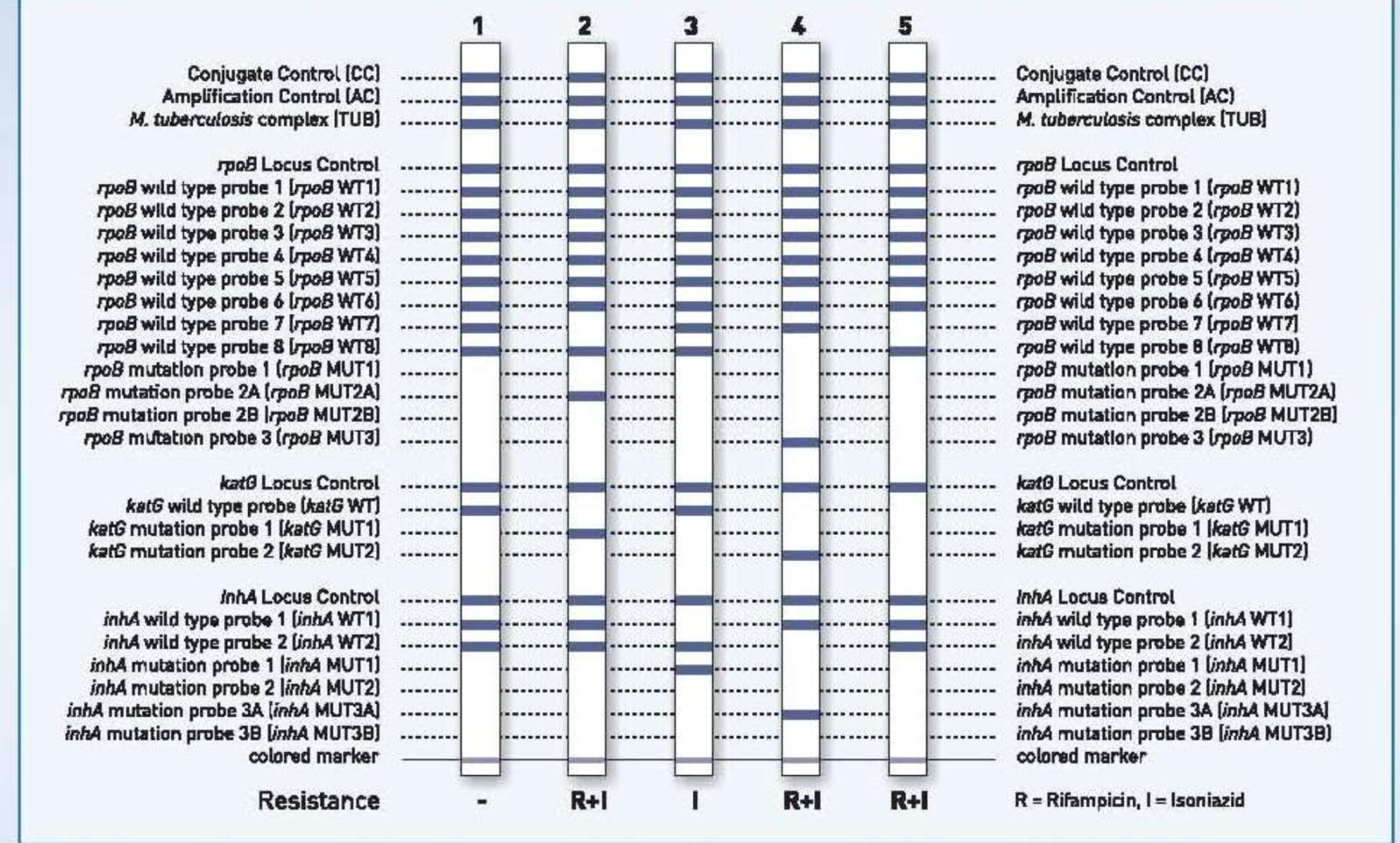
Identification of *M.tuberculosis* complex and its resistance to Rifampicin and/or Isoniazid using the Genotype MTBDRplus. Interpretation of the Line Probe Assay (Adapted[30])

### DST Testing

Drug susceptibility testing was performed on baseline isolates and samples from patients suspected of treatment failure or relapse in order to identify the presence of resistance to any of the TB drugs under study. Drug susceptibility testing was performed as previously described [20]. Briefly, the *M. tuberculosis* confirmed culture was added to a liquid medium (BACTEC MGIT 960, Becton Dickenson) with 1 % proportion test of susceptibility to isoniazid 0.2 μg/ml, rifampin 1 μg/ml, streptomycin 2 μg/ml, ethambutol 5 μg/ml, kanamycin 5 μg/ml, and ofloxacin 4 μg/ml. For one to be classified as MDR-TB infected they had to be resistant to isoniazid and rifampicin (whether or not they had resistance to other anti-TB drugs). Each batch consisted of one H37RV sensitivity strain which was run for quality control purpose.

### Statistical analysis

To determine associations with the primary outcome: MDR-TB, univariate and multivariate logistic regression was employed. A cut-off p-value of 0.05 was used to determine the strength of associations. Chi-squared test was used to compare the distributions of test results for LPA and DST. All the results were presented in tables. Agreements between LPA method and the reference method (DST) were assessed using Kappa statistics. Cohen’s kappa values were interpreted as follows: from 0.00 to 0.20 indicating slight agreement; from 0.21 to 0.40 suggest fair agreement; from 0.41 to 0.6 show moderate agreement; from 0.61 to 0.80 indicate substantial agreement; and above 0.8 show excellent agreement [21] Only available data was considered in the analysis of the different variables (only complete cases were considered depending on the variables being analysed. R statistical language was used to analyse the data [22].

## Results

A total of 128 *M. tuberculosis* positive samples from Lilongwe site were run on molecular Genotype MTBDRplus VER 2.0 LPA and compared with phenotypic DST whereas for the Blantyre site only LPA was conducted. The median age of the participants was 35 (IQR: 29.0 – 70.0) for Lilongwe site, while for the Blantyre site the median age was 21 (IQR: 11.0 – 38.25)

Overall the incidence of MDR-TB by Genotype MTBDRplus VER 2.0 LPA was found to be 18/128 (14.06%) whereas that by DST was 30/128 (23.44%). RIF Mono-resistance (*rpoB* gene mutation) was found to occur at 8/128 (6.25%) for LPA and 8/128 (6.25%) for DST, while a large number of samples were found be non-resistant to all drugs tested with 102/128 (79.69%) for LPA and 90/128 (70.31%) for DST. The distributions of the test results under LPA and DST did not differ significantly (Chi-squared test: df = 4, p-value = 0.1991) (Table 1).

**Table 1.**
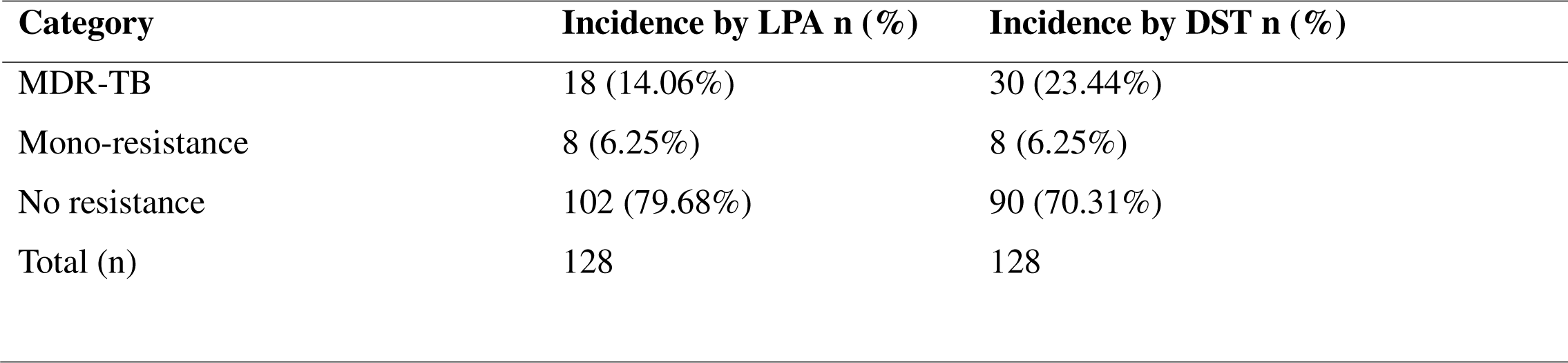
Comparison of the incidence of MDR-TB by LPA vs DST.

Next we cross-tabulated the results from the two tests in order to determine the agreement between the two tests with the row being DST and the column the Genotype MTBDRplus VER 2.0 LPA. We found significant agreement between Genotype MTBDRplus VER 2.0 LPA and DST in detecting MDR-TB (Table 2). A kappa statistic of 0.75 (95% CI: 0.62-0.88) was obtained. Using MTBDRplus VER 2.0 LPA (compared to DST) at the Lilongwe site, the sensitivity, specificity, positive predicting value, and negative predicting value for detection of mono-resistance were 50%, 97%, 50%, and 97% respectively; and for detection of multi-drug resistance the sensitivity, specificity, positive predicting value, and negative predicting value were 62%, 100%, 100% and 90% respectively (Table 3).

**Table 2.** Agreement of the incidence of MDR-TB by LPA vs DST.

| **Genotype MTBDRplus VER 2.0 LPA** |  |  |  |  |
| --- | --- | --- | --- | --- |
|  |  | MDR | Mono-resistance | No resistance |
| **DST** | MDR | 18 | 4 | 8 |
|  | Mono-resistance | 0 | 4 | 4 |
|  | No resistance | 0 | 0 | 90 |

**Table 3.**
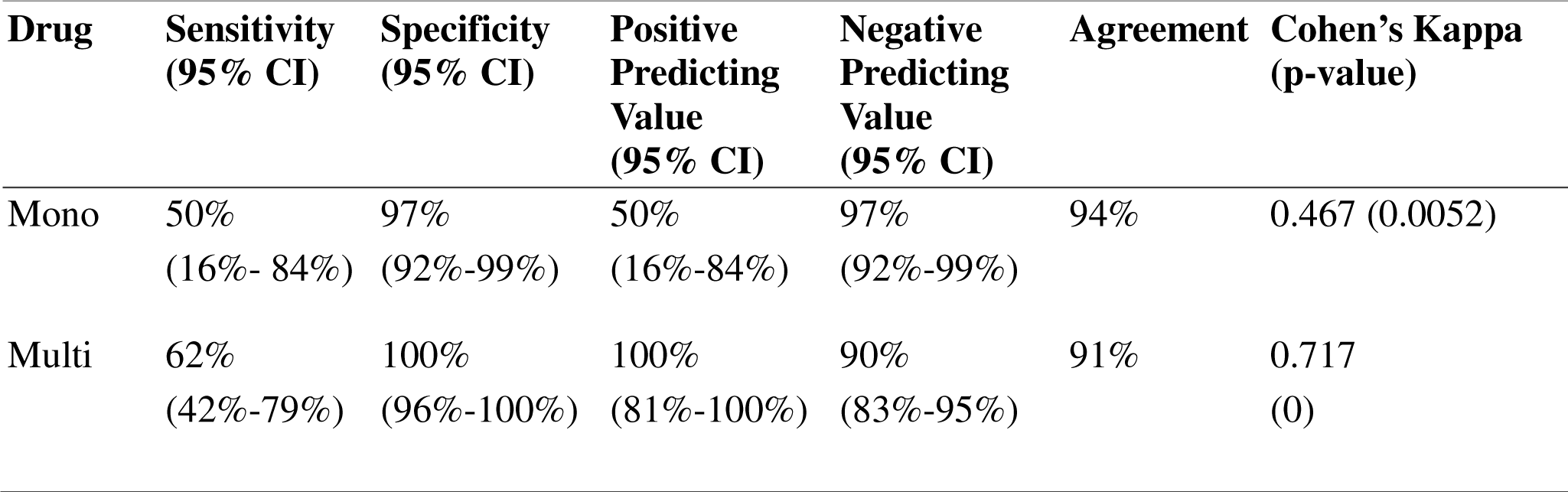
Performance characteristics of MTBDRplus VER 2.0 LPA when compared to DST method in detection of mono-resistance (RIF resistance), and multi-drug resistance.

To determine the incidence of MDR-TB at the Lilongwe site only, a kappa statistical analysis was conducted and found to be 67.08% with a 95% confidence interval of 51.74% - 82.42%. Using MTBDRplus VER 2.0 LPA (compared to DST) at the Lilongwe site, the sensitivity, specificity, positive predicting value, and negative predicting value for detection of RIF resistance were 66%, 100%, 100%, and 87% respectively; for detection of INH resistance the sensitivity, specificity, positive predicting value, and negative predicting value were 17%, 100%, 100% and 80% respectively; while for multi-drug resistance the sensitivity, specificity, positive predicting value, and negative predicting value were 100% each (Table 3).

To determine associations with the primary outcome: MDR-TB, univariate and multivariate logistic regression was employed and a cut-off p-value of 0.05 was used to determine the strength of associations. There was no significant difference in the likelihood of developing MDR-TB between those less than 36 years old, and those older. The likelihood of developing MDR-TB was lower for males compared to females (odds ratio was 0.23, and 95% CI was 0.076-0.646). No significant difference was observed in the likelihood of developing MDR-TB between participants with smear grades of scanty, 1+, 2+, and 3+, and those that were negative (Table 4).

**Table 4.** Logistic regression analysis by age, sex and smear grade.

| **Variable** | **Stratum** | **Frequency**<br>**(%)** | **COR**<br>**(95% CI)** | **P-value** | **AOR**<br>**(95% CI)** | **P-value** |
| --- | --- | --- | --- | --- | --- | --- |
| **Age** | < 36<br>years | 56<br>(60.2%) | 1 |  | 1 |  |
|  | >= 36<br>years | 37<br>(39.8%) | 0.74<br>(0.28, 1.9) | 0.5397 | 0.95<br>(0.33, 2.74) | 0.9311 |
| **Sex** | Female | 42 (42%) | 1 |  | 1 |  |
|  | Male | 58 (58%) | 0.28<br>(0.1, 0.69) | 0.0073<br>9 | 0.27<br>(0.09, 0.74) | 0.0125 |
| **Smear**<br>**grade** | Negative | 34<br>(36. 6%) | 1 |  | 1 |  |
|  | Scanty | 10<br>(10.8%) | 1.91<br>(0.38, 9.57) | 0.4184 | 2.52<br>(0.46, 14.45) | 0.2833 |
|  | 1+ | 17<br>(18.3%) | 0.64<br>(0.15, 2.34) | 0.5105 | 0.91<br>(0.2, 3.84) | 0.8983 |
|  | 2+ | 18<br>(19.4%) | 0.44<br>(0.09, 1.73) | 0.2685 | 0.56<br>(0.1, 2.4) | 0.4526 |
|  | 3+ | 14<br>(15.1%) | 0.76<br>(0.18, 2.89) | 0.6996 | 0.95<br>(0.21, 3.96) | 0.9478 |

## Discussion

This is the first study to describe the burden of MDR-TB based on Genotype MTBDRplus VER 2.0 LPA and phenotypic DST results in Malawi. Although other studies have reported DR-TB in Malawi, the data were only based on INH mono-resistance [19] or limited to only a single urban hospital [23]. Additionally, this is the first laboratory based study to combine MDR-TB data from two different regions of the country.

Both LPA and Cartridge Based Nucleic Acid Amplification Test (NAAT), have been endorsed by the World Health Organization (WHO), but there is no clarity regarding the superiority of one over the other[13]. This notwithstanding, studies elsewhere have demonstrated that LPAs can significantly reduce the time to treatment initiation for MDR-TB patients [13,24]. In the current study, for both Lilongwe and Blantyre sites, all samples 128 (100%) and 30 (100%) respectively had a positive TUB band on LPA suggesting they were all members of *M. tuberculosis* complex. The sensitivity of LPAs in detecting *M. tuberculosis* complex has been questioned in some studies owing to the fact that it has been unable to detect the TUB band correctly in up to 6% of smear positive *M. tuberculosis* samples [13].

Out of 128 samples from Lilongwe LPA was able to detect 14.06% as MDR-TB compared to 23.44% by DST representing 61% concordance. These results are inconsistent with previous studies [14] where LPA was able to detect 66 out the 68 (97%) DST MDR-TB isolates correctly. RIF mono-resistance was found to occur at 6.06% and 7.07% for Genotype MTBDRplus VER 2.0 LPA and DST respectively (Table 1) whereas no INH mono-resistance was detected. The high rates of RIF mono-resistance which is usually rare could be attributed to the fact that over 70% of TB cases in Malawi were found in HIV infected individuals at the height of the pandemic[25,26]. In the current study we were unable to conduct a comparative analysis of HIV status owing to a small sample size. Interestingly, the WHO policy is that LPAs may not completely replace mycobacterial culture and DST [13] but studies continue to suggest the increasing importance of LPAs as it has shown to significantly reduce time to treatment initiation for MDR-TB patients. The sensitivities of LPA compared to DST for the detection of RIF, INH and MDR-TB were 66%, 17% and 100% respectively whereas the specificities were 100% throughout (Table 3). In our results the sensitivity of LPA for RIF and INH resistance were lower compared to other studies [13,14,27]. Although INH sensitivity was detected at a lower level, other studies have suggested correlation with RIF resistance [27]. Additionally other studies in India [28] and Germany[15] have reported very high sensitivities and specificities for INH resistance detection by LPA. Our failure to detect INH mono-resistance could in part due to the fact that 10% to 25% of INH resistant strains are thought to have mutations outside *katG* and *inhA* loci [12,29]. Several other factors have been proposed for low sensitivity LPA results including reagent quality, preparation and presence of PCR inhibitors [14]. Overall our results suggest that Genotype MTBDRplus VER 2.0 LPA may be as good as DST at detecting MDR-TB consistent with previous studies that have reported higher sensitivities and specificities of LPA [12].

At the Blantyre site (KUHeS/MLW TB laboratory), only LPA was conducted on a total number of 30 *M. tuberculosis* positive samples. We could not conduct DST owing to inadequate infrastructure. LPA was able to detect 1/30 (3.3%) as being INH mono-resistant in which none were neither RIF nor MDR-TB. The study population had HIV status as a variable, but due to small sample size, we did not calculate the association between HIV and development of MDR-TB owing to the absence of any MDR-TB. At 95% confidence interval, the point estimate for MDR-TB was 0% while for mono-resistance TB was 3.3%.

In this study, the majority of the suspected (52.5%) and lab-confirmed (56%) MDR-TB patients were from the age group between 15 and 45 years in concordance with the national data. The high frequencies of MDR-TB among young age groups may indicate the possibility of propagation of MDR-TB in the community because of the higher mobility of youth.

Our results show that there is a positive association between MDR-TB and Sex (Table 4). The likelihood of males developing MDR-TB was lower compared to females with odds ratio of 28% (95% CI is 1% - 78%). However, this may be attributed to the fact that most males do not present to the hospital when they get ill and may not truly represent what obtains in the community.

In conclusion, this study has demonstrated that MTBDRplus VER 2.0 LPA can perform better at detecting incidence of MDR-TB. Combing DST and LPA results would remain the best option in detecting MDR TB among patients.

This study was limited by the fact that we were unable to conduct a complete comparison of the two methods for the Blantyre isolates. Additionally, using secondary data meant we were unable to collect some social-demographic information such as occupation, HIV status (especially at National TB Reference Laboratory), previous TB treatment, residence, and antibiotic history. Therefore, the final multivariable logistic regression model was not properly adjusted for due to this missing information which could lead to significant residual confounding.

## Conclusion

There is high incidence of MDR-TB among patients whose samples are sent to Lilongwe National TB Reference Laboratory which otherwise remain undetected. Larger studies covering the three geo-political regions of the country need to be undertaken. With performance characteristics compared to other conventional methods of detecting MDR-TB, a short turnaround time to diagnosis, and the ability to simultaneously detect rifampicin and isoniazid resistance, makes LPA an excellent and reliable tool for the early detection of multidrug-resistant tuberculosis. LPA should be implemented in all regions of Malawi for detection MDR-TB in addition to DST.

## Data Availability

All data produced in the present study are available upon reasonable request to the authors

## Acknowledgement

We would like to acknowledge the Helse Nord Tuberculosis Initiative (HNTI) for funding this study.

## Authors’ contributions

VN, MN and TS conceived the study. VN, TS, DC, SD performed the investigations. MN, TK, TS HT and AM analyzed the data. VN, MN, TS, TK, AM, MN, BB and YK wrote the report. MN, VN and TS oversaw the research. All authors contributed to the study design and reviewed the report.

